# The contribution of common regulatory and protein-coding *TYR* variants in the genetic architecture of albinism

**DOI:** 10.1101/2021.11.01.21265733

**Authors:** Vincent Michaud, Eulalie Lasseaux, David J. Green, Dave T. Gerrard, Claudio Plaisant, UK Biobank Eye and Vision Consortium, Tomas Fitzgerald, Ewan Birney, Benoît Arveiler, Graeme C. Black, Panagiotis I. Sergouniotis

## Abstract

Genetic diseases have been historically segregated into rare Mendelian and common complex conditions.^1,2^ Large-scale studies using genome sequencing are eroding this distinction and are gradually unmasking the underlying complexity of human traits.^3–8^ Aiming to gain insights into the genetic architecture of rare recessive disorders, we studied a cohort of 1,313 individuals with albinism, an archetypal Mendelian condition. We investigated the contribution of protein-coding and regulatory variants both rare and common. We focused on *TYR*, the gene encoding tyrosinase, and found that a high-frequency promoter variant, *TYR* c.-301C>T [rs4547091], modulates the penetrance of a prevalent, disease-associated missense change, *TYR* c.1205G>A [rs1126809]. We also found that homozygosity for a haplotype formed by three common, functionally-relevant variants, *TYR* c.[-301C;575C>A;1205G>A], is associated with a high probability of receiving an albinism diagnosis (OR>82). This genotype is also associated with reduced visual acuity and increased central retinal thickness in UK Biobank participants. Finally, we report how the combined analysis of rare and common variants increases diagnostic yield and informs genetic counselling in families with albinism.

## MAIN

There is abundant evidence supporting the view that rare genetic diseases are caused by rare, high-impact variants in individual genes. However, for most known rare disorders, it is not possible to identify such pathogenic changes in every affected proband, leaving significant diagnostic and knowledge gaps.^9–11^ In recent years, the emergence of comprehensive rare disease and population-based resources that link genomic and phenotypic data (e.g. UK Biobank^12^, Genomics England 100,000 Genomes Project^13^) has offered unprecedented opportunities for genetic discovery. Through integrative analysis of these datasets, we can now achieve line-of-sight for uncovering complex molecular explanations in people with rare disorders who have hitherto remained undiagnosable.

Albinism, a rare condition characterised by decreased ocular pigmentation and altered visual system organisation^14^, had a pivotal role in the study of human genetics tracing back to the early 20^th^ century.^15,16^ At least 20 genes are now known to be associated with this disorder and the current diagnostic yield of genetic testing in affected cohorts approaches 75%.^17–19^ Most people with a molecular diagnosis of albinism carry biallelic variants in *TYR*, the gene encoding the rate-limiting enzyme of melanin biosynthesis^20^. Building on recent work^17,21^, we sought to increase our understanding of the genetic complexity of this archetypal disorder.

A cohort of 1208 people with albinism underwent testing of ≤19 albinism-related genes; these individuals were not known to be related and had predominantly European ancestries (Supplementary Table 1). A further 105 probands with albinism were identified in the Genomics England 100,000 Genomes Project dataset^13^. A ‘control’ cohort of 29,497 unrelated individuals that had no recorded diagnosis/features of albinism was also identified in this resource (Fig.1, Supplementary Table 2, Methods).

**Figure 1.**
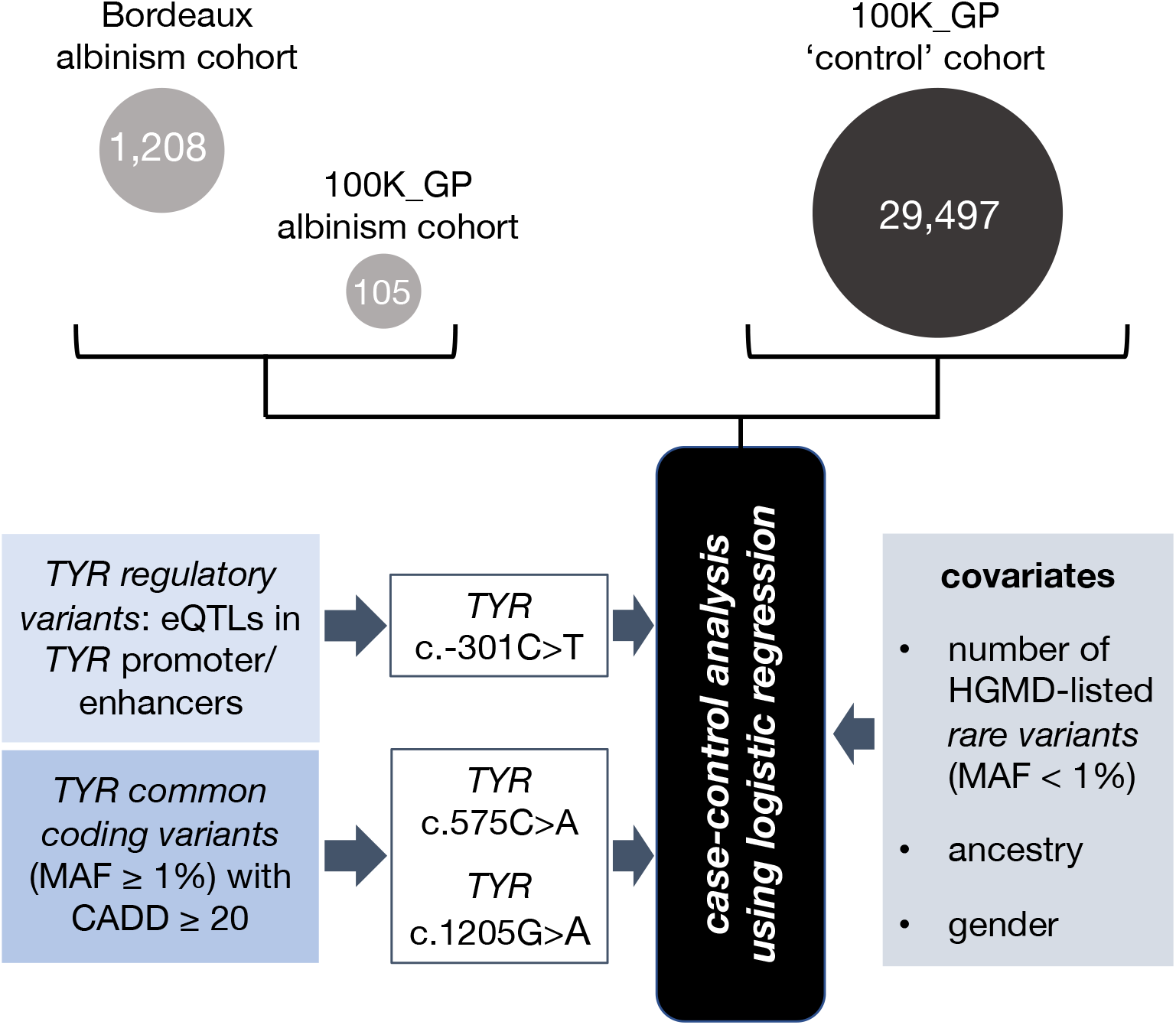
Outline of the case-control study design A case-control analysis was performed to gain insights into the contribution of protein-coding and regulatory variation, at the common and rare ends of the allele-frequency spectrum, in albinism. The majority of participants in the ‘case’ cohort (1208/1313) were identified through the database of the University Hospital of Bordeaux Molecular Genetics Laboratory, France. All these probands had at least one key ocular feature of albinism, *i.e*. nystagmus or prominent foveal hypoplasia. The remaining 105/1313 cases were identified through the Genomics England 100,000 Genomes Project dataset and had a diagnosis of albinism or a phenotype deemed consistent with partial/ocular albinism. The ‘control’ cohort included 29,497 unrelated individuals from the Genomics England 100,000 Genomes Project dataset, none of whom had a recorded diagnosis of albinism. Genotypes that include selected *TYR* haplotypes in homozygous state were studied. Haplotypes of interest were defined as those formed by combinations of *TYR* variants that are predicted to be functionally relevant; three variants met the pre-determined criteria set for regulatory (*TYR* c.-301C>T) and common protein-coding (*TYR* c.575C>A and c.1205G>A) variants. The associated haplotypic blocks were analysed further using logistic regression (see Methods). 100K_GP, Genomics England 100,000 Genomes Project; eQTLs, expression quantitative trait loci; MAF, minor allele frequency; CADD, Combined Annotation Dependent Depletion score; HGMD, Human Gene Mutation Database v2021.2. *TYR* variant numbering is based on the transcript with the following identifiers: NM_000372.5 and ENST00000263321.6.

To gain insights into the contribution of common variants to the genetic architecture of albinism, we studied the impact of protein-coding changes that have minor allele frequency [MAF] ≥1% and are predicted by a computational algorithm to be functionally relevant (CADD score^22^ ≥20). For *TYR*, two such variants were identified: c.575C>A (p.Ser192Tyr) [rs1042602] and c.1205G>A (p.Arg402Gln) [rs1126809]. Multiple associations have been recorded for these two changes including skin/hair pigmentation (for both variants), macular thickness (for c.575C>A) and iris colour (for c.1205G>A).^23^ Furthermore, each of these changes has been shown to decrease TYR enzymatic activity *in vitro*.^24,25^ Importantly, there is evidence suggesting that c.1205G>A is acting as a hypomorphic variant and is causing a mild form of albinism when in compound heterozygous state with a complete loss-of-function *TYR* mutation.^26^ It is also noted that the MAF of this variant in European populations is around 27% and that multiple unaffected homozygous individuals have been reported (including >2,000 people in the control subset of the Genome Aggregation Database [gnomAD] v2.1.1)^27^.

To gain insights into the contribution of regulatory variants, we studied the impact of changes that alter *TYR* regulatory elements (*i.e*. the *TYR* promoter or ENCODE-listed enhancers)^28^ and affect *TYR* gene expression (*i.e*. they are known *TYR* expression quantitative trait loci [*eQTL*]). One such variant was identified, c.-301C>T [rs4547091], a fetal retinal pigment epithelium (RPE) selective eQTL^29^. This change is known to alter a binding site for the transcription factor *OTX2* in the *TYR* promoter, and the reference allele (c.-301C) has been shown to lead to a remarkable decrease in promoter activity *in vitro*^30^.

Focusing on individual sequence alterations without consideration for variant interactions and/or patterns of linkage disequilibrium can lead to masking of complex underlying mechanisms. To overcome this pitfall, we avoided an independent analysis of each of the *TYR* c.-301C>T, c.575C>A and c.1205G>A changes and instead studied the haplotype blocks that they form. Eight possible haplotypes [2^3^] and 36 possible haplotype pairs [2^3-1^ x (2^3^+1)] may be encountered. We focused only on the 8 haplotype pairs that include homozygous alleles (Fig.2A) for two reasons: (1) in homozygous individuals, the underlying haplotypes can be unambiguously determined, even in cases where segregation/phasing data are unavailable; (2) in autosomal recessive disorders like *TYR*-related albinism, phenotypic abnormalities are the result of the combined effect of two alleles; by analyzing only homozygous cases, the effect of a specific haplotype can be isolated and estimated with greater precision.

We used Firth regression analysis^31,32^ to study how *TYR* haplotypes in homozygous state affect the risk of albinism (i.e. the probability of having a diagnosis of albinism). This increasingly recognised logistic regression approach has been designed to handle small, imbalanced datasets (which are common in studies of rare conditions) and allows for adjustment of key covariates (which is not possible in contingency table methods) (Fig.1, Methods). The results are shown in Fig.2B and Supplementary Table 3. This analysis identified a number of pertinent points that are discussed below.

**Figure 2.**
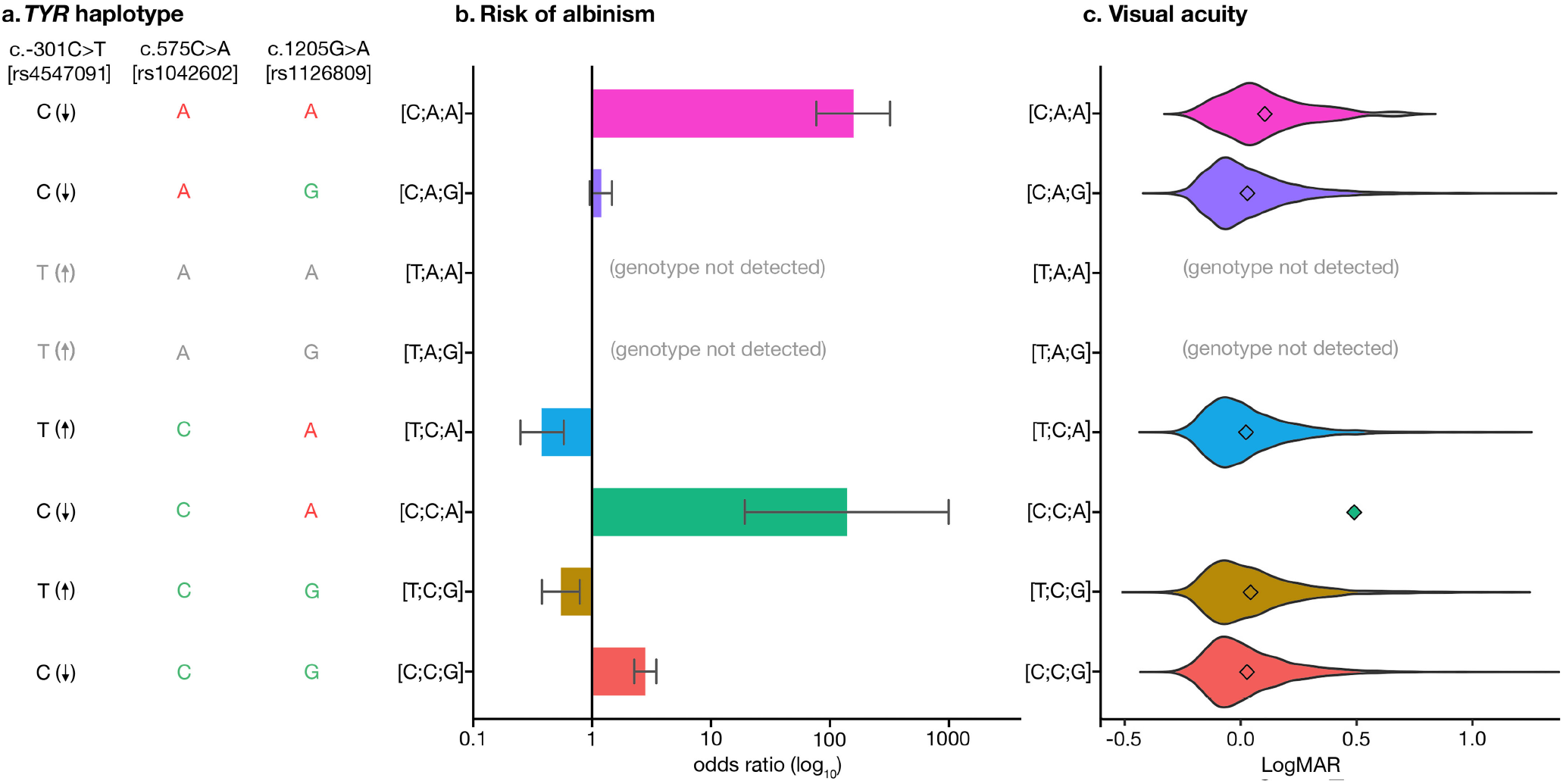
Common *TYR* variants form haplotypes that affect risk of albinism and visual performance. **a**. The *TYR* haplotypes that were studied are shown. The reference allele of the *TYR* c.-301C>T [rs4547091] promoter variant reduces gene expression and is shown as C (↓); the non-reference allele increases gene expression and is shown as T (↑). The reference alleles of the c.575C>A (p.Ser192Tyr) [rs1042602] and the c.1205G>A (p.Arg402Gln) [rs1126809] missense variants are highlighted in green font while the non-reference alleles are highlighted in red font. As no homozygotes for the *TYR* [-301T;575A;1205A] and [-301T;575A;1205G] haplotypes were detected, these combinations are highlighted in grey font. **b**. Risk of albinism (i.e. probability of receiving a diagnosis of albinism) in people carrying selected *TYR* haplotypes in homozygous state. Odds ratio >1 suggests an increased risk while odds ratio <1 suggests a decreased risk. Further information including numeric data can be found in Supplementary Table 3. **c**. Visual acuity in UK Biobank participants carrying selected *TYR* haplotypes in homozygous state. Vision near 0.0 LogMAR is considered normal while vision >0.5 LogMAR is considered moderate/severe visual impairment. The Kruskal-Wallis p-value was 8 × 10^−11^. Further information including numeric data can be found in Supplementary Table 5.

We found that the penetrance of the hypomorphic *TYR* c.1205G>A variant^26^ is modulated by the *TYR* c.-301C>T promoter change. When c.1205G>A is encountered in a homozygous state and in combination with the c.-301C allele of the promoter variant (which reduces *TYR* expression), the risk of albinism is high (OR>24; see [C;A;A] and [C;C;A] in Fig.2B). In contrast, homozygosity for c.1205G>A combined with the c.-301T allele (which increases *TYR* expression) has a protective effect (OR<0.7; see [T;C;A] in Fig.2B). This observation is in keeping with previous studies suggesting that penetrance can be modified by the joint functional effects of regulatory and protein-coding variants.^33^ We here provide a key illustration of this mechanism in the context of a recessively-acting hypomorphic variant.

Alongside this, we found that homozygosity for the *TYR* c.-301C>T promoter variant protects against albinism (OR 0.3-0.7; see [T;C;A] and [T;C;G] in Fig.2B). Notably, the allele frequency of the protective c.-301T allele, approaches 80% in people of African ancestries and is around 40% in people of European ancestries^27^ (see Supplementary Fig.1 for the geographical distribution of the associated variants/haplotypes). It can be speculated that variation in this *TYR* promoter position partly accounts for the relatively low prevalence of *TYR*-related albinism in people of African ancestries (Supplementary Table 4).

Our findings also highlight that homozygosity for the haplotype formed by the c.-301C allele of the promoter variant (which reduces *TYR* expression) and the non-reference alleles of the two common missense changes, c.575C>A and c.1205G>A, is associated with a significant increase in the risk of albinism (OR >82; see [C;A;A] in Fig.2B). This haplotype is present in ∼1% of people with European ancestries in the 1000 Genomes Project (phase 3)^34^, and the associated risk of albinism is comparable to that of a loss-of-function Mendelian mutation.

When the *TYR* c.[-301C;575A;1205A] and c.[-301C;575C;1205A] haplotypes (corresponding to [C;A;A] and [C;C;A] in Fig.2B) were factored in as Mendelian variants in a clinical-grade analysis of the case cohort, the diagnostic yield increased from 57% (692/1208) to 76% (916/1208) (Figure 3). It is noted that current genetic laboratory pipelines are generally suboptimally set up to identify these complex high-risk haplotypes, especially when filtering is based on the rarity of individual variants.

**Figure 3.**
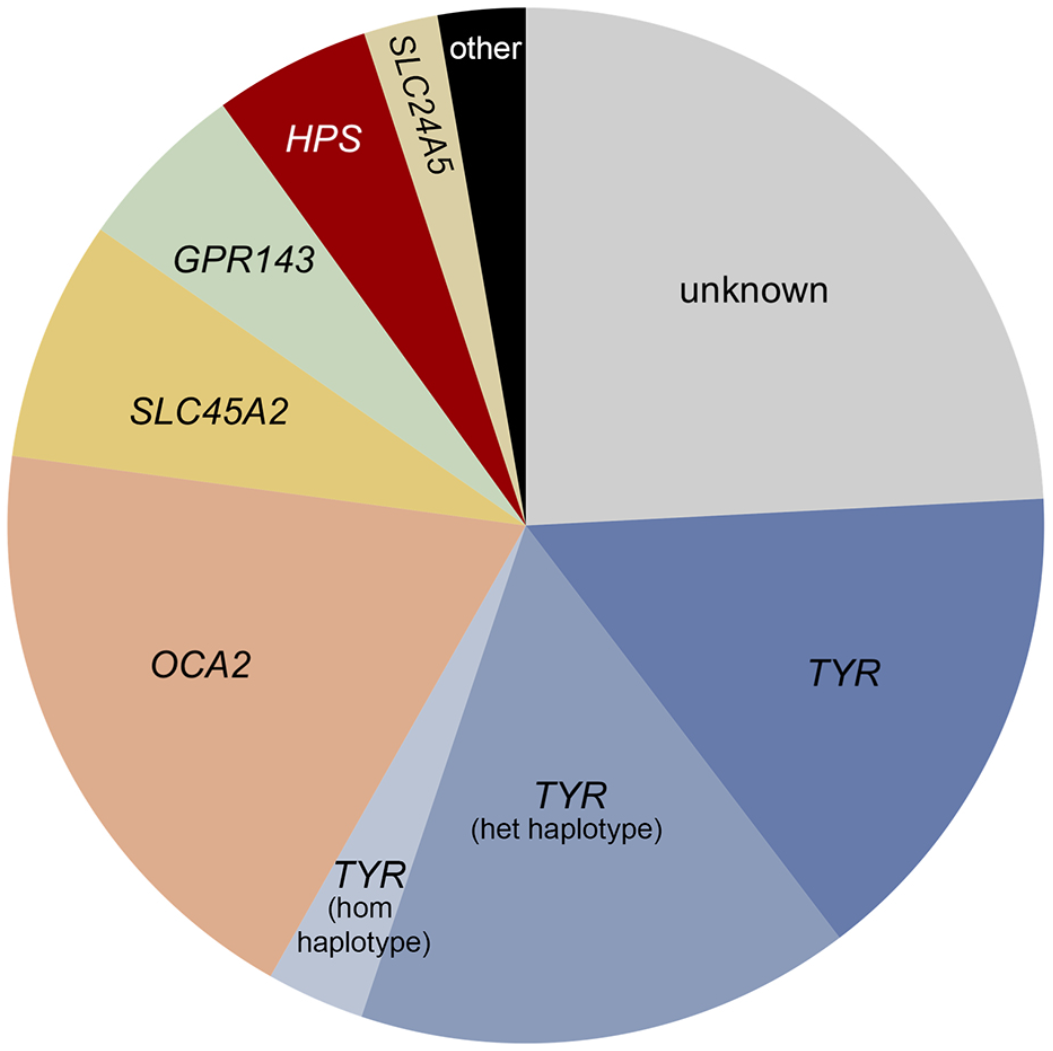
High level molecular diagnoses in 1208 probands from the University Hospital of Bordeaux albinism cohort. It was not possible to detect a molecular diagnosis in 24% of cases (“unknown” category). The following genes were implicated in the remaining probands: *TYR* (34%), *OCA2* (19%), *SLC45A2* (8%), *GPR143* (5%), Hermansky-Pudlak syndrome (HPS) related genes (5%), *SLC24A5* (2%), other albinism-related genes (3%). A significant subset of cases with *TYR*-related albinism were found/presumed to carry either the *TYR* c.[-301C;575A;1205A] or the *TYR* c.[-301C;575C;1205A] haplotype (15% in heterozygous state [“*TYR* het haplotype” category]; 3% in homozygous state [“*TYR* hom haplotype” category]). Further information including a list with all molecular diagnoses can be found in Supplementary Table 1.

We subsequently studied the impact of the *TYR* c.[-301C;575A;1205A] haplotype (corresponding to [C;A;A] in Fig.2B) in UK Biobank participants. We found that people who were homozygous for this haplotype had, on average, reduced visual acuity (mean LogMAR vision 0.10; Kruskal-Wallis p-value 8 × 10^−11^ with all pairwise comparisons involving [C;A;A] being statistically significant; Fig.2C and Supplementary Table 5). As visual acuity is a quantitative endophenotype of albinism, this finding provides additional evidence supporting the functional significance of this complex haplotype. A similar trend was noted when central retinal thickness, another albinism endophenotype, was assessed (Kruskal-Wallis p-value < 2 × 10^−16^; Supplementary Fig.2 and Supplementary Table 6). We expect that future studies analysing visual function and ocular structure in this group of homozygous individuals will provide key insights into the elusive link between RPE melanin synthesis and visual system organisation.^35^ Furthermore, we anticipate that the study of cellular models specific to these homozygous cases (e.g. human induced pluripotent stem cell-derived RPE) will advance our understanding of the molecular pathology of albinism.

Lastly, we quantified the risk of albinism associated with combinations of rare and common variants. For each study participant, we estimated two key contributors to an individual’s risk. First, we counted the number of rare, presumed Mendelian variants in albinism-related genes; single nucleotide variants that have MAF<1% and are labelled as disease-causing (DM) in the Human Gene Mutation Database (HGMD) v2021.2^36^ were considered. Subsequently, we counted the number of common “risk genotypes” in *TYR* (*i.e*. c.-301C, c.575A and/or c.1205A in heterozygous state). We found that the presence of >4 common *TYR* risk genotypes confers an increased risk of albinism even in the absence of a rare, HGMD-listed variant (OR>3.6; Table 1). We also found that, when a single heterozygous HGMD-listed variant co-occurs with >1 common *TYR* risk genotype, the risk of albinism is increased (OR>4.2 for rare variants in any albinism-related gene, OR>1.2 for rare variants in *TYR;* Table 1). These observations provide a basis for more precise genetic counseling in families with albinism.

**Table 1.** Contribution of different classes of albinism-related variants to disease risk

| combination of common<sup>a</sup><br>and rare<sup>b</sup> risk genotypes | odds<br>ratio<sup>c</sup> | 95% confidence<br>interval | p-value |
| --- | --- | --- | --- |
| 6 common + 0 rare | 391 | 165 - 982 | 0.000016 |
| 5 common + 0 rare | 7.8 | 3.6 - 16.5 | <0.00001 |
| 4 common + 0 rare | 3.4 | 1.9 - 6.1 | 0.000014 |
| 3 common + 0 rare | 1.5 | 0.8 - 2.6 | 0.17 |
| 2 common + 0 rare | 2.3 | 1.3 - 4 | 0.002 |
| 1 common + 0 rare | 1.1 | 0.6 - 1.9 | 0.84 |
| 0 common + 0 rare | 0.1 | 0.1 - 0.2 | <0.00001 |
| 6 common + 1 rare<sup>d</sup> | 217 | 26.7 - 2519 | 0.00001 |
| 5 common + 1 rare | 302 | 156 - 566 | <0.00001 |
| 4 common + 1 rare | 38 | 22 - 69 | <0.00001 |
| 3 common + 1 rare | 8.7 | 4.9 - 15.6 | <0.00001 |
| 2 common + 1 rare | 7.5 | 4.2 - 13.5 | <0.00001 |
| 1 common + 1 rare | 6 | 3.2 - 11.2 | <0.00001 |
| 0 common + 1 rare | 2.6 | 1.2 - 5.3 | 0.016 |
| 0 common + 2 rare | 163 | 75 - 367 | <0.00001 |
| 6 common + 1 rare *TYR*<sup>d</sup> | 120 | 6 - 17919 | 0.003 |
| 5 common + 1 rare *TYR* | 178 | 82 - 425 | <0.00001 |
| 4 common + 1 rare *TYR* | 38.6 | 20 - 80 | <0.00001 |
| 3 common + 1 rare *TYR* | 5.3 | 2.6 - 11 | <0.00001 |
| 2 common + 1 rare *TYR* | 2.4 | 1.2 - 5.4 | 0.02 |
| 1 common + 1 rare *TYR*<sup>d</sup> | 0.6 | 0.2 - 1.9 | 0.43 |
| 0 common + 1 rare *TYR*<sup>d</sup> | 2.3 | 0.4 - 8 | 0.28 |
| 0 common + 2 rare *TYR*<sup>d</sup> | 446 | 100 - 4270 | <0.00001 |

<sup>a</sup> presence of each of the following *TYR* variants in heterozygous state was considered as one common risk genotype: c.-301C [rs4547091]; c.575C>A (p.Ser192Tyr) [rs1042602]; c.1205G>A (p.Arg402Gln) [rs1126809]. *TYR* variant numbering is based on the transcript with the following identifiers: NM\_000372.5 and ENST00000263321.6.
<sup>b</sup> autosomal albinism-related genes were inspected and each heterozygous protein-coding change with a minor-allele frequency <1% and a “disease-causing” (DM) label in the Human Gene Mutation Database (HGMD) v2021.2 was considered as one rare risk genotype. Such genotypes in the *TYR* gene are labelled “rare *TYR*”.
<sup>c</sup> Firth regression analysis with gender and ancestry as covariates was used to estimate odds ratios and p-values; 1,121 probands with albinism and 29,453 unrelated controls were included (Supplementary Table 1).
<sup>d</sup> Less than 10 individuals were included in the smallest subgroup of these categories, reducing the reliability of the associated findings.

One potential limitation of this study is our inability to stringently match the albinism cases with the unaffected controls, especially in terms of recent ancestry (which can be correlated with skin pigmentation). Although ancestry was included as a parameter in our regression model, this analysis was imperfect as it was not possible to reliably assign genetic ancestry to most albinism cases. It is known that inability to fully account for differences in ancestral background between cases and controls can lead to false-positive association signals.^37^ We used a combination of orthogonal approaches to evaluate the robustness and generalisability of our findings. First, we used 35 presumed neutral single-nucleotide variants to calculate the genomic inflation factor lambda (λGC)^38,39^; λ_median_ was found to be 1.04, in keeping with limited confounding by ancestry (Supplementary Table 7). Subsequently, we performed targeted sub-analyses of the available cohorts; the results of three focused case-control studies supported our key findings and increased confidence in the validity of the detected associations (Supplementary Figure 3 and Supplementary Tables 8-10).

In conclusion, we have shown that a significant proportion of albinism risk arises from genetic susceptibility linked to common variants. Furthermore, our findings suggest that rare and common protein-coding variation in *TYR* should be considered in the context of regulatory haplotypes. The concepts discussed here are highly likely to be relevant to the understanding of other rare disorders, and haplotype-based approaches are expected to narrow the diagnostic gap for significant numbers of patients. Future work will embrace more diverse populations and focus on integrating both common and rare variants (including single-nucleotide and copy-number changes) into a single genetic risk score at scale.

## ONLINE METHODS

### Cohort characteristics and genotyping

#### University Hospital of Bordeaux albinism cohort

Individuals with albinism were identified through the database of the University Hospital of Bordeaux Molecular Genetics Laboratory, France. This is a national reference laboratory that has been performing genetic testing for albinism since 2003 and has been receiving samples from individuals predominantly based in France (or French-administered overseas territories). Information on the dermatological and ophthalmological phenotypes was available and all people included in the study had at least one of the key ocular features of albinism, *i.e*. nystagmus or absence of a foveal pit (prominent foveal hypoplasia). No pre-screening based on genotype was undertaken and only individuals who were not knowingly related were included.

Genetic testing, bioinformatic analyses, and clinical interpretation were performed as previously described.^17,26^ Briefly, most participants had gene-panel testing of 19 genes associated with albinism (*TYR, OCA2, TYRP1, SLC45A2, SLC24A5, C10ORF11, GPR143, HPS 1* to 10, *LYST, SLC38A8*) using IonTorrent platforms. High-resolution array-CGH (comparative genomic hybridization) was also used to detect copy number variants in these 19 genes. All genetic changes of interest were confirmed with an alternative method (e.g. Sanger sequencing or quantitative PCR). Clinical interpretation of variants was performed using criteria consistent with the 2015 American College of Medical Genetics and Genomics (ACMG) best practice guidelines^40^. Generally, variants with MAF ≥1% in large publicly available datasets (e.g. gnomAD^27^) were considered unlikely to be disease-causing. We note that the genetic findings in a subset of this cohort (70%; 845/1208) have been partly reported in a previous publication^17^ (see Supplementary Table 1 for further information).

Due to the limited number of genes screened in this cohort, it was not possible to reliably assess genetic ancestry and to objectively assign individuals to ancestry groups. Attempting to mitigate this, we processed available data on self-identified ethnicity that were collected through questionnaires. Responses were inspected and stratification into five broad continental groups (European, African, Admixed American, East Asian, South Asian) was performed.

Informed consent was obtained from all participants or their parents in the case of minors. The study was approved by the relevant local ethics committee (Comité de Protection des Personnes Sud-Ouest et Outre Mer III, Bordeaux, France) and all investigations were conducted in accordance with the tenets of the Declaration of Helsinki.

#### Genomics England 100,000 Genomes Project cohort

Clinical and genomic data from the Genomics England 100,000 Genomes Project were accessed through a secure Research Environment that is available to registered users. This dataset was collected as part of a national genome sequencing initiative.^41^ Enrolment was coordinated by Genomics England Limited and participants were recruited mainly at National Health Service (NHS) Hospitals in the UK.^13^ Clinical information was recorded in Human Phenotype Ontology (HPO)^42^ terms and International Classification of Diseases (ICD) codes. Genome sequencing was performed in DNA samples from 78,195 individuals using Illumina HiSeq X systems (150 base-pair paired-end format). Reads were aligned using the iSAAC Aligner v03.16.02.19 and small variants were called using Starling v2.4.7.^43^ Aggregation of single-sample gVCFs was performed using the Illumina software gVCF genotyper v2019.02.29; normalisation/decomposition was implemented by vt version 0.57721^44^. The multi-sample VCF was then split into 1,371 roughly equal chunks to allow faster processing and the loci of interest were queried using bcftools v1.9^45^ (see https://research-help.genomicsengland.co.uk/display/GERE/ for further information). Only variants that passed all provided site quality control criteria were processed. In addition, we filtered out genotypes with: genotype score <20; read depth <10; allele balance <0.2 and >0.8 for heterozygotes; allele balance >0.1 or <0.9 for homozygotes (reference and alternate, respectively). Genomic annotation was performed using Ensembl VEP^46^; one additional annotation was included - presence of a variant in HGMD v2021.2^36^ with a “disease-causing” (DM) label.

Ancestry inference was performed in this cohort using principal component analysis. Data from the 1000 genomes project (phase 3) dataset^41^ was used and five broad super-populations were projected (European, African, Admixed American, East Asian, South Asian) (further information on this can be found online at https://research-help.genomicsengland.co.uk/display/GERE/Ancestry+inference).

We focused on a pre-determined subset of the Genomics England 100,000 Genomes Project dataset that includes only unrelated probands (n=29,602). 105 of these individuals had a diagnosis of albinism, *i.e*. the ICD-10 term “Albinism” [E70.3] and/or the HPO terms “Albinism” [HP:0001022], “Partial albinism” [HP:0007443] or “Ocular albinism” [HP:0001107] were assigned. Together with the University Hospital of Bordeaux cases, these 105 probands formed the ‘case’ cohort (for the albinism risk analysis). The remaining 29,497 probands had no recorded diagnosis or phenotypic features of albinism and formed the ‘control’ cohort. We note that in-depth ophthalmic phenotyping was not routinely undertaken in Genomics England 100,000 Genomes Project participants. Thus, we cannot be certain that a small number of individuals with mild/subclinical forms of albinism are not included in the control cohort.

### Identifying functional regulatory and protein-coding variants

#### Regulatory variants

Focusing on *TYR*, we identified changes that are likely to have an impact on gene regulation by selecting variants that:

- are known *TYR* eQTLs.
- alter *TYR* cis-regulatory elements, including the promoter of the gene.

To identify eQTLs, we inspected the eQTL catalogue^47^ and used data from the Genotype-Tissue Expression v8 (GTEx)^48^ and Eye Genotype Expression (EyeGEx)^49^ projects. To identify regulatory elements, we used the ENCODE 3 (ENCyclopedia Of DNA Elements phase 3) dataset; the SCREEN (Search Candidate cis-Regulatory Elements by ENCODE) v10 interface was utilised to query this resource for regions flagged as candidate cis-regulatory elements (see https://screen.encodeproject.org/ for further information and definitions).^28^ Additional putative regulatory elements were identified by inspecting chromatin accessibility peaks in RPE samples in DESCARTES (the Developmental Single Cell Atlas of Gene Regulation and Expression)^50^ and through an extensive search of the biomedical literature (e.g.^29^). All these queries were conducted in January 2021.

#### Common protein-coding variants

Focusing on *TYR*, we identified common changes that are likely to have an impact on protein function by selecting variants that:

- have a CADD PHRED-scaled score ≥20. CADD is a widely-used integrative annotation tool built from more than 60 genomic features. A PHRED-scaled score ≥10 indicates a raw score in the top 10% of all possible single nucleotide variants, while a score ≥20 indicates a raw score in the top 1%;^22^ it is noted that a cut-off of 20 has balanced sensitivity and specificity (90% and 69% respectively)^51^ in the context of this non-diagnostic setting.
- alter protein-coding sequences - including missense changes, nonsense variants and small insertions/deletions; variants with a potential role on splicing (e.g. synonymous changes and variants altering splice donor/acceptor sites) were not included.
- have “total” MAF ≥1% in gnomAD v2.1.1^27^

#### Rare protein-coding variants

Focusing on 19 albinism-related genes *(TYR, OCA2, TYRP1, SLC45A2, SLC24A5, C10ORF11, GPR143, HPS 1* to *10, LYST, SLC38A8*), we identified rare changes that are likely to have an impact on protein function by selecting variants that:

- are labelled as disease-causing (DM) in HGMD v2021.2.
- are included in the following HGMD v2021.2 “mutation type” categories: missense/nonsense, splicing, small deletions, small insertions or small indels; gross deletions, gross insertions/duplications and complex rearrangements were not analysed.
- have “total” MAF <1% in gnomAD v2.1.1^27^.

### Case-control analysis to estimate albinism risk

The effect of homozygosity for selected *TYR* haplotypes (formed by one common regulatory change, c.-301C>T, and two common protein-coding variants, c.575C>A and c.1205G>A variants) on albinism risk (i.e. the probability of receiving a diagnosis of albinism) was estimated using data from the University Hospital of Bordeaux albinism cohort and the Genomics England 100,000 Genomes Project dataset. A case-control analysis of a binary trait (presence/absence of albinism) was conducted assuming a recessive model. Logistic regression using Firth’s bias reduction method^31,32^ was utilised (as implemented in “logistf” R package)^50^. The following covariates were included: gender, number of rare HGMD-listed variants and ancestry (Supplementary Table 3).

Although ancestry was included as a co-variate in our logistic regression model, this analysis was imperfect as it was not possible to objectively determine genetic ancestry in the University Hospital of Bordeaux albinism cohort (as mentioned above, self-identified ethnicity was instead used as a surrogate). Confounding by ancestry (i.e population stratification) is therefore a possibility. This can arise if there are systematic differences in ancestral background between cases and controls; these differences can result in significantly different allele/genotype frequencies between the compared groups that may lead to spurious association signals.^52^ We attempted to quantify the bias in our data by calculating the genomic inflation factor lambda (λGC). λGC is conceptually simple and involves using a set of random genetic markers to quantitatively estimate the structural differences between the case and control populations.^38,39^ The selected λGC markers have to be unlinked and should not be expected to show an association with the trait under study (albinism or skin pigmentation in this case). We therefore selected 35 single-nucleotide variants that (i) had a CADD PHRED-scaled score <5 (*i.e*. were unlikely to be functionally relevant) and (ii) were genotyped both by the gene panels used in the University Hospital of Bordeaux albinism cohort and the genome sequencing assays used in Genomics England 100,000 Genomes Project participants (Supplementary Table 7). Subsequently, case-control comparisons were made for each of these 35 λGC markers using Firth regression analysis. The resulting test statistics were then used to calculate the median value of λGC.

To further understand the impact of recent ancestry on our results, we analysed selected subsets of the case and control cohorts. First, we performed sub-analysis of the Genomics England 100,000 Genomes Project cases (n=105) and controls (n=29,497). Then we aimed to compare groups that were matched both in terms of genetic ancestry and geographical origin; thus we focused on the Genomics England 100,000 Genomes Project cases (n=76) and controls (n=22,927) that have European ancestries (as inferred by principal component analysis). Finally, we repeated our primary analysis using individuals from both the University Hospital of Bordeaux and the Genomics England 100,000 Genomes Project cohorts but this time focusing only on the cases (n=1,107) and controls (n=22,927) that have European ancestries. The results are shown in Supplementary Figure 3 and Supplementary Tables 8-10.

### Analysis of visual acuity and foveal thickness in UK Biobank participants

The effect of homozygosity for selected *TYR* haplotypes was studied in UK Biobank participants. UK Biobank is a biomedical resource containing in-depth genetic and health information from >500,000 individuals from across the UK^12^. A subset of UK Biobank volunteers underwent enhanced phenotyping including visual acuity testing (131,985 individuals) and imaging of the central retina (84,748 individuals)^53^; the latter was obtained using optical coherence tomography (OCT), a non-invasive imaging test that rapidly generates cross-sectional retinal scans at micrometre-resolution.^54^ All UK Biobank volunteers analysed as part of this study were imaged using the 3D OCT-1000 Mark II device (Topcon, Japan); the relevant methodology has been previously described.^53^ Notably, only 24 UK Biobank participants are assigned a diagnosis of albinism (data field 41270; ICD-10 term “Albinism” [E70.3]) of which only 7 had visual acuity measurements and none had OCT imaging; 19 additional individuals had a diagnosis of albinism in their primary care record data (resource 591). Given that reduced visual acuity and increased central retinal thickness (due to underdevelopment of the fovea) are two key hallmark features of albinism we investigated the impact of *TYR* risk haplotypes on these quantitative endophenotypes.

First, genotyping array data were used to obtain genotypes for *TYR* c.575C>A [rs1042602] and *TYR* c.1205G>A [rs1126809] (data field 22418 including information from the Applied Biosystems UK Biobank Axiom Array containing 825,927 markers). In contrast to these two changes, the *TYR* c.-301C>T [rs4547091] variant was not directly captured by the array. However, high-quality (>99.9%) imputation data on this promoter change were available (data field 22828).

Subsequently, we calculated the mean of the right and left LogMAR visual acuity for each UK Biobank volunteer (data fields 5201 and 5208, “instance 0” datasets). These visual acuity measurements were subsequently used to compare visual performance between groups of people with different homozygous haplotype combinations. As the obtained distributions deviated from normality (Fig.2C), the Kruskal-Wallis test was used. Pair-wise comparisons were performed and the p-values were adjusted using the Benjamini-Hochberg method (Supplementary Table 5).

To obtain central foveal thickness measurements from UK Biobank OCT images, we calculated the mean of the right and left central retinal thickness (defined as the average distance between the hyperreflective bands corresponding to the RPE and the internal limiting membrane (ILM), across the central 1 mm diameter circle of the ETDRS grid) for each UK Biobank volunteer).^51^ The obtained measurements were then subsequently used to compare central macular thickness between groups of UK Biobank volunteers with different homozygous *TYR* haplotype combinations. As some of the obtained distributions deviated from normality (Supplementary Fig.2), the Kruskal-Wallis test was used. Pair-wise comparisons were performed and the p-values were adjusted using the Benjamini-Hochberg method (Supplementary Table 6).

## Data Availability

All data produced in the present study are available upon reasonable request to the authors. Phenotypic and genomic data from the Genomics England 100,000 Genomes Project and the UK Biobank project can be accessed by registered users with relevant approved applications; details on this can be found online at https://www.genomicsengland.co.uk/about-gecip/for-gecip-members/data-and-data-access and https://www.ukbiobank.ac.uk respectively.

## DATA AND CODE AVAILABILITY STATEMENT

Genomics England 100,000 Genomes Project data are available through an access procedure described at https://www.genomicsengland.co.uk/about-gecip/for-gecip-members/data-and-data-access. UK Biobank data are available through a procedure described at http://www.ukbiobank.ac.uk/using-the-resource/. All other data supporting the findings of this study are available within the article (including its supplementary information files) or from the authors upon request. The scripts used to analyse the datasets included in this study can be found at https://github.com/davidjohngreen.

## ACKNOWLEDGEMENTS

We acknowledge the following sources of funding: the Wellcome Trust (200990/Z/16/Z, Transforming Genetic Medicine Initiative); Christopher Green; Retina UK and Fight for Sight (GR586, RP Genome Project - UK Inherited Retinal Disease Consortium); Health Education England; UK National Institute for Health Research (NIHR) Clinical Lecturer Programme (CL-2017-06-001); the French Albinism Association (Genespoir).

This research was made possible through access to the data and findings generated by the 100,000 Genomes Project. The 100,000 Genomes Project is managed by Genomics England Limited (a wholly owned company of the Department of Health and Social Care). The 100,000 Genomes Project is funded by the National Institute for Health Research and NHS England. The Wellcome Trust, Cancer Research UK and the Medical Research Council have also funded research infrastructure. The 100,000 Genomes Project uses data provided by patients and collected by the National Health Service as part of their care and support.

We acknowledge the contribution of the Genomics England Research Consortium to the 100,000 Genomes Project. Members of this Consortium include: John C. Ambrose, Prabhu Arumugam, Roel Bevers, Marta Bleda, Freya Boardman-Pretty, Christopher R. Boustred, Helen Brittain, Mark J. Caulfield, Georgia C. Chan, Greg Elgar, Tom Fowler, Adam Giess, Angela Hamblin, Shirley Henderson, Tim J. P. Hubbard, Rob Jackson, Louise J. Jones, Dalia Kasperaviciute, Melis Kayikci, Athanasios Kousathanas, Lea Lahnstein, Sarah E. A. Leigh, Ivonne U. S. Leong, Javier F. Lopez, Fiona Maleady-Crowe, Meriel McEntagart, Federico Minneci, Loukas Moutsianas, Michael Mueller, Nirupa Murugaesu, Anna C. Need, Peter O’Donovan, Chris A. Odhams, Christine Patch, Mariana Buongermino Pereira, Daniel Perez-Gil, John Pullinger, Tahrima Rahim, Augusto Rendon, Tim Rogers, Kevin Savage, Kushmita Sawant, Richard H. Scott, Afshan Siddiq, Alexander Sieghart, Samuel C. Smith, Alona Sosinsky, Alexander Stuckey, Mélanie Tanguy, Ana Lisa Taylor Tavares, Ellen R. A. Thomas, Simon R. Thompson, Arianna Tucci, Matthew J. Welland, Eleanor Williams, Katarzyna Witkowska, Suzanne M. Wood

This research was conducted using the UK Biobank Resource under projects 53144, 49978 and 2112. Members of the UK Biobank Eye and Vision Consortium include: Tariq Aslam, Sarah Barman, Jenny Barrett, Paul Bishop, Catey Bunce, Roxana Carare, Usha Chakravarthy, Michelle Chan, Valentina Cipriani, Alexander Day, Parul Desai, Bal Dhillon, Andrew Dick, Cathy Egan, Sarah Ennis, Paul Foster, Marcus Fruttiger, John Gallacher, David Garway-Heath, Jane Gibson, Dan Gore, Jeremy Guggenheim, Chris Hammond, Alison Hardcastle, Simon Harding, Ruth Hogg, Pirro Hysi, Pearse A. Keane, Peng T. Khaw, Anthony Khawaja, Gerassimos Lascaratos, Andrew J. Lotery, Phil Luthert, Tom Macgillivray, Sarah Mackie, Bernadette Mcguinness, Gareth Mckay, Martin Mckibbin, Danny Mitry, Tony Moore, James Morgan, Zaynah Muthy, Eoin O’Sullivan, Chris Owen, Praveen Patel, Euan Paterson, Tunde Peto, Axel Petzold, Jugnoo Rahi, Alicja Rudnicka, Jay Self, Sobha Sivaprasad, David Steel, Irene Stratton, Nicholas Strouthidis, Cathie Sudlow, Caroline Thaung, Dhanes Thomas, Emanuele Trucco, Adnan Tufail, Veronique Vitart, Stephen Vernon, Ananth Viswanathan, Cathy Williams, Katie Williams, Jayne Woodside, Max Yates, Jennifer Yip, Yalin Zheng, Robyn Tapp, Denize Atan, Alexander Doney, Naomi Allen, Thomas Littlejohns, Panagiotis Sergouniotis, Graeme Black.

Lastly, we acknowledge the help of Cécile Courdier at the University Hospital of Bordeaux Molecular Genetics Laboratory and of Jamie Ellingford at the University of Manchester.

## REFERENCES

1. Claussnitzer, M. et al. A brief history of human disease genetics. Nature 577, 179–189 (2020).

2. Shendure, J., Findlay, G. M. & Snyder, M. W. Genomic Medicine–Progress, Pitfalls, and Promise. Cell 177, 45–57 (2019).

3. Niemi, M. E. K. et al. Common genetic variants contribute to risk of rare severe neurodevelopmental disorders. Nature 562, 268–271 (2018).

4. Tilghman, J. M. et al. Molecular genetic anatomy and risk profile of Hirschsprung’s disease. N. Engl. J. Med. 380, 1421–1432 (2019).

5. Turro, E. et al. Whole-genome sequencing of patients with rare diseases in a nationalhealth system. Nature 583, 96 (2020).

6. Chung, B. H. Y., Chau, J. F. T. & Wong, G. K.-S. Rare versus common diseases: a false dichotomy in precision medicine. NPJ genomic Med. 6, (2021).

7. Khan, M. et al. Resolving the dark matter of ABCA4 for 1054 Stargardt disease probands through integrated genomics and transcriptomics. Genet. Med. 22, 1235–1246 (2020).

8. Harper, A. R. et al. Common genetic variants and modifiable risk factors underpin hypertrophic cardiomyopathy susceptibility and expressivity. Nat. Genet. 2021 532 53, 135– 142 (2021).

9. Boycott, K. M. et al. International cooperation to enable the diagnosis of all rare genetic diseases. Am. J. Hum. Genet. 100, 695 (2017).

10. Ferreira, C. R. The burden of rare diseases. Am. J. Med. Genet. A 179, 885–892 (2019).

11. Chakravarti, A. Magnitude of Mendelian versus complex inheritance of rare disorders. Am. J. Med. Genet. A (2021). doi:10.1002/AJMG.A.62463

12. Bycroft, C. et al. The UK Biobank resource with deep phenotyping and genomic data. Nature 562, 203–209 (2018).

13. Genomics England. The National Genomic Research Library v5.1. (2019). Available at: https://doi.org/10.6084/m9.figshare.4530893.v6.

14. Kruijt, C. C. et al. The phenotypic spectrum of albinism. Ophthalmology 125, 1953–1960 (2018).

15. Farabee, W. C. Notes on Negro Albinism. Science 17, 75 (1903).

16. Garrod, A. E. The Croonian Lectures on Inborn Errors of Metabolism. Lancet 172, 1–7 (1908).

17. Lasseaux, E. et al. Molecular characterization of a series of 990 index patients with albinism. Pigment Cell Melanoma Res. 31, 466–474 (2018).

18. Wang, C. et al. [Spectrum of pathological genetic variants among 405 Chinese pedigrees affected with oculocutaneous albinism]. Chin J Med Genet 37, 725–730 (2020).

19. Mauri, L. et al. Clinical evaluation and molecular screening of a large consecutive series of albino patients. J. Hum. Genet. 62, 277–290 (2017).

20. Pavan, W. J. & Sturm, R. A. The genetics of human skin and hair pigmentation. Annu. Rev. Genomics Hum. Genet. 20, 41–72 (2019).

21. Campbell, P. et al. Clinical and genetic variability in children with partial albinism. Sci. Rep. 9, 16576 (2019).

22. Rentzsch, P., Witten, D., Cooper, G. M., Shendure, J. & Kircher, M. CADD: Predicting the deleteriousness of variants throughout the human genome. Nucleic Acids Res. 47, D886– D894 (2019).

23. Buniello, A. et al. The NHGRI-EBI GWAS Catalog of published genome-wide association studies, targeted arrays and summary statistics 2019. Nucleic Acids Res. 47, D1005 (2019).

24. Chaki, M. et al. Molecular and functional studies of tyrosinase variants among Indian oculocutaneous albinism type 1 patients. J. Invest. Dermatol. 131, 260–262 (2011).

25. Jagirdar, K. et al. Molecular analysis of common polymorphisms within the human Tyrosinase locus and genetic association with pigmentation traits. Pigment Cell Melanoma Res. 27, 552– 564 (2014).

26. Monfermé, S. et al. Mild form of oculocutaneous albinism type 1: phenotypic analysis of compound heterozygous patients with the R402Q variant of the TYR gene. Br. J. Ophthalmol. 103, 1239–1247 (2019).

27. Karczewski, K. J. et al. The mutational constraint spectrum quantified from variation in 141,456 humans. Nature 581, 434–443 (2020).

28. The ENCODE Project Consortium et al. Expanded encyclopaedias of DNA elements in the human and mouse genomes. Nature 583, 699–710 (2020).

29. Liu, B. et al. Genetic analyses of human fetal retinal pigment epithelium gene expression suggest ocular disease mechanisms. Commun. Biol. 2, 186 (2019).

30. Reinisalo, M., Putula, J., Mannermaa, E., Urtti, A. & Honkakoski, P. Regulation of the human tyrosinase gene in retinal pigment epithelium cells: The significance of transcription factor orthodenticle homeobox 2 and its polymorphic binding site. Mol. Vis. 18, 38–54 (2012).

31. Heinze, G. & Schemper, M. A solution to the problem of separation in logistic regression. Stat. Med. 21, 2409–2419 (2002).

32. Firth, D. Bias Reduction of Maximum Likelihood Estimates. Biometrika 80, 27 (1993).

33. Castel, S. E. et al. Modified penetrance of coding variants by cis-regulatory variation contributes to disease risk. Nat. Genet. 50, 1327 (2018).

34. Auton, A. et al. A global reference for human genetic variation. Nat. 2015 5267571 526, 68–74 (2015).

35. Mason, C. & Guillery, R. Conversations with Ray Guillery on albinism: linking Siamese cat visual pathway connectivity to mouse retinal development. Eur. J. Neurosci. 49, 913–927 (2019).

36. Stenson, P. D. et al. The Human Gene Mutation Database (HGMD ®): optimizing its use in a clinical diagnostic or research setting. Hum. Genet. 139, 1197–1207 (2020).

37. Freedman, M. L. et al. Assessing the impact of population stratification on genetic association studies. Nat. Genet. 2004 364 36, 388–393 (2004).

38. Devlin, B. & Roeder, K. Genomic control for association studies. Biometrics 55, 997–1004 (1999).

39. Dadd, T., Weale, M. E. & Lewis, C. M. A critical evaluation of genomic control methods for genetic association studies. Genet. Epidemiol. 33, 290–298 (2009).

40. Richards, S. et al. Standards and guidelines for the interpretation of sequence variants: a joint consensus recommendation of the American College of Medical Genetics and Genomics and the Association for Molecular Pathology. Genet. Med. 2015 175 17, 405–423 (2015).

41. Turnbull, C. et al. The 100 000 Genomes Project: bringing whole genome sequencing to the NHS. BMJ 361, 1687 (2018).

42. Köhler, S. et al. The Human Phenotype Ontology in 2021. Nucleic Acids Res. 49, D1207– D1217 (2021).

43. Raczy, C. et al. Isaac: ultra-fast whole-genome secondary analysis on Illumina sequencing platforms. Bioinformatics 29, 2041–2043 (2013).

44. Tan, A., Abecasis, G. R. & Kang, H. M. Unified representation of genetic variants. Bioinformatics 31, 2202–2204 (2015).

45. Danecek, P. et al. Twelve years of SAMtools and BCFtools. Gigascience 10, giab008 (2021).

46. McLaren, W. et al. The Ensembl Variant Effect Predictor. Genome Biol. 17, (2016).

47. Nurlan, K. et al. A compendium of uniformly processed human gene expression and splicing quantitative trait loci. Nat. Genet. 53, 1290–1299 (2021).

48. The GTEx Consortium. The GTEx Consortium atlas of genetic regulatory effects across human tissues. Science 369, 1318–1330 (2020).

49. Ratnapriya, R. et al. Retinal transcriptome and eQTL analyses identify genes associated with age-related macular degeneration. Nat. Genet. 51, 606–610 (2019).

50. Domcke, S. et al. A human cell atlas of fetal chromatin accessibility. Science 370, (2020).

51. van der Velde, K. J. et al. GAVIN: Gene-Aware Variant INterpretation for medical sequencing Genome Biol. 18, 1–10 (2017).

52. Marchini, J., Cardon, L. R., Phillips, M. S. & Donnelly, P. The effects of human population structure on large genetic association studies. Nat. Genet. 2004 365 36, 512–517 (2004).

53. Chua, S. Y. L. et al. Cohort profile: design and methods in the eye and vision consortium of UK Biobank. BMJ Open 9, e025077 (2019).

54. Drexler, W. & Fujimoto, J. G. State-of-the-art retinal optical coherence tomography. Prog. Retin. Eye Res. 27, 45–88 (2008).

55. Currant, H. et al. Genetic variation affects morphological retinal phenotypes extracted from UK Biobank optical coherence tomography images. PLoS Genet. 17, e1009497 (2021).

