## Supplementary Information for "The contribution of common regulatory and protein-coding *TYR* variants in the genetic architecture of albinism"

### SUPPLEMENTARY FIGURES

**Supplementary Figure 1**. Geographic distribution of selected *TYR* common variants and haplotypes.

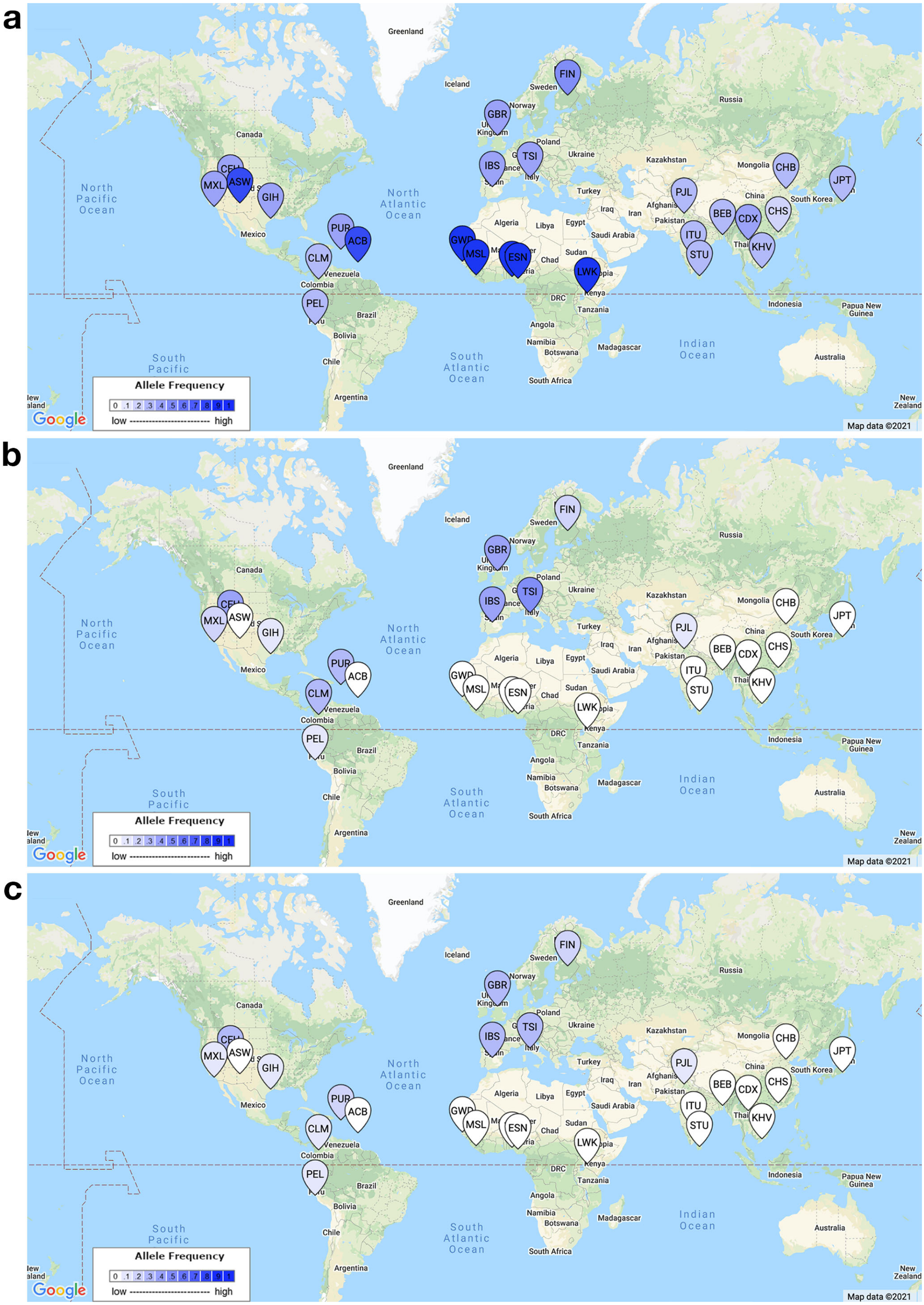

**d**

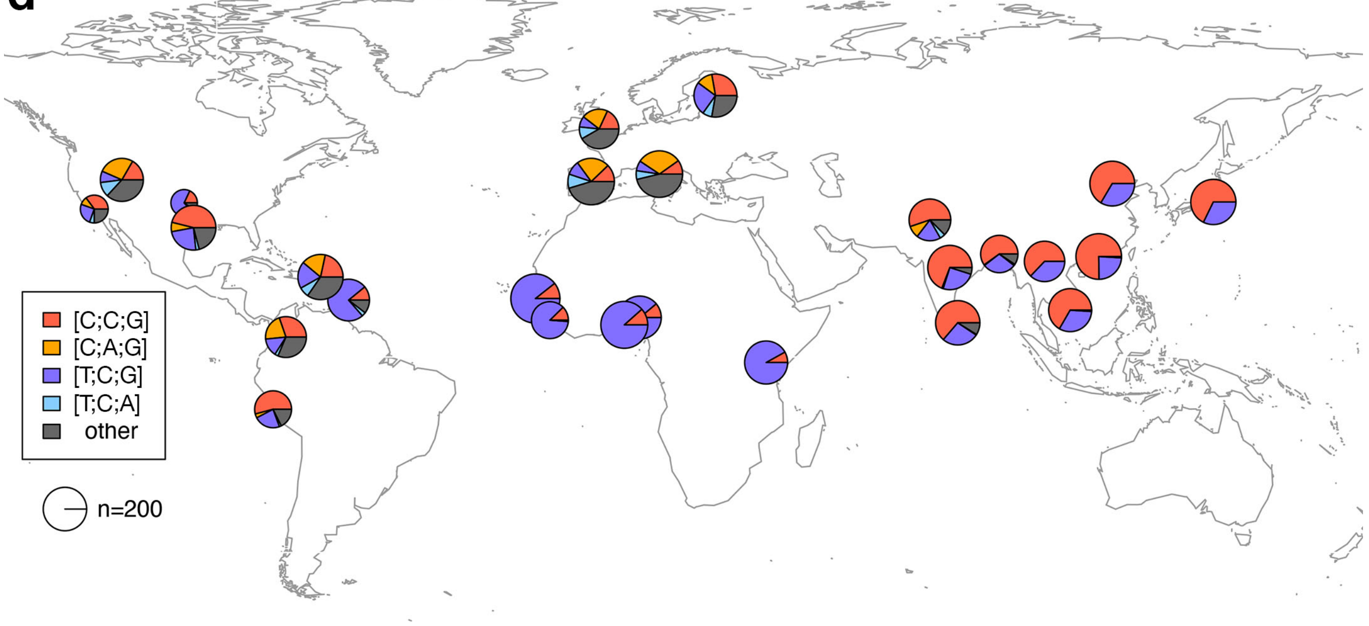


**a-c**. Geographic distribution of allele frequencies for the **(a)** *TYR* c.-301C>T [rs4547091], **(b)** *TYR* c.575C>A (p.Ser192Tyr) [rs1042602], and **(c)** *TYR* c.1205G>A (p.Arg402Gln) [rs1126809] variants across 1000 Genomes Project (phase 3) populations[1]. The LDpop tool[2] (LDlink 5.1 release) was used to generate these maps; numeric data on sample size and allele frequencies are available at https://ldlink.nci.nih.gov/?tab=ldpop. The allele frequencies of these variants across relevant subpopulations in the Simons Genome Diversity Project dataset[3] were comparable; these can be found in the Atlas of Variant Age resource (https://human.genome.dating/)[4]. This tool also provided insights into the estimated age of these changes:

- c.-301C>T [rs4547091]: T is predicted to be the ancestral allele and the estimated age of the derived C allele using a joint clock model was >1,000,000 years. This is a low-quality estimate that might be imprecise (quality score 0.2; values near 1 indicate high quality and values near 0 indicate low quality).
- c.575C>A [rs1042602]: C is the ancestral allele and the estimated age of the derived A allele using a joint clock model was ~23,000 years (quality score 0.9).
- c.1205G>A [rs1126809]: G is the ancestral allele and the estimated age of the derived A allele using a joint clock model was ~33,000 years (quality score 0.6).

Notably, the phylogeographic patterns for c.575C>A and c.1205G>A have been previously studied using a Bayesian coalescent approach; the estimated ages for c.575C>A and c.1205G>A were ~15,600 and ~29,400 years respectively.[5]

**d.** Geographic distribution of haplotype frequencies for the *TYR* c.[-301C;575C;1205G], c.[-301C;575A;1205G], c.[-301T;575C;1205G] and c.[-301T;575C;1205A] haplotypes (corresponding to [C;C;G], [C;A;G], [T;C;G] and [T;C;A] respectively) across 1000 Genomes Project (phase 3) populations[1]. The category 'other' contains two types of haplotypes: (i) 728 (out of a total of 5008; 15%) haplotypes from individuals who are heterozygous at two or more of the three variant positions (phase could not be inferred in these cases), and (ii) two further confirmed haplotypes, c.[-301C;575A;1205A] and c.[-301C;575C;1205A]), that were counted only twelve and two times, respectively. The R packages *maps*[6] and *plotrix*[7] were used in generating this map. Population information and geographic coordinates were obtained from www.internationalgenome.org.

*AFR (African)*: ACB (African Caribbean in Barbados); ASW (African Ancestry in Southwest US); ESN (Esan in Nigeria), GWD (Gambian in Western Division, The Gambia); LWK (Luhya in Webuye, Kenya); MSL (Mende in Sierra Leone); YRI (Yoruba in Ibadan, Nigeria).

*AMR (American)*: CLM (Colombian in Medellin, Colombia); MXL (Mexican Ancestry in Los Angeles, California); PEL (Peruvian in Lima, Peru); PUR (Puerto Rican in Puerto Rico).

*EAS (East Asian)*: CDX (Chinese Dai in Xishuangbanna, China); CHB (Han Chinese in Bejing, China); CHS (Southern Han Chinese); JPT (Japanese in Tokyo, Japan); KHV (Kinh in Ho Chi Minh City, Vietnam).

*EUR (European)*: CEU (Utah Residents with Northern and Western European ancestry); FIN (Finnish in Finland); GBR (British in England and Scotland); IBS (Iberian populations in Spain); TSI (Toscani in Italy).

*SAS (South Asian)*: BEB (Bengali in Bangladesh); GIH (Gujarati Indian from Houston, Texas); ITU (Indian Telugu in the United Kingdom); PJL (Punjabi in Lahore, Pakistan); STU (Sri Lankan Tamil in the United Kingdom).

**Supplementary Figure 2**. Impact of common *TYR* variants on central retinal thickness.

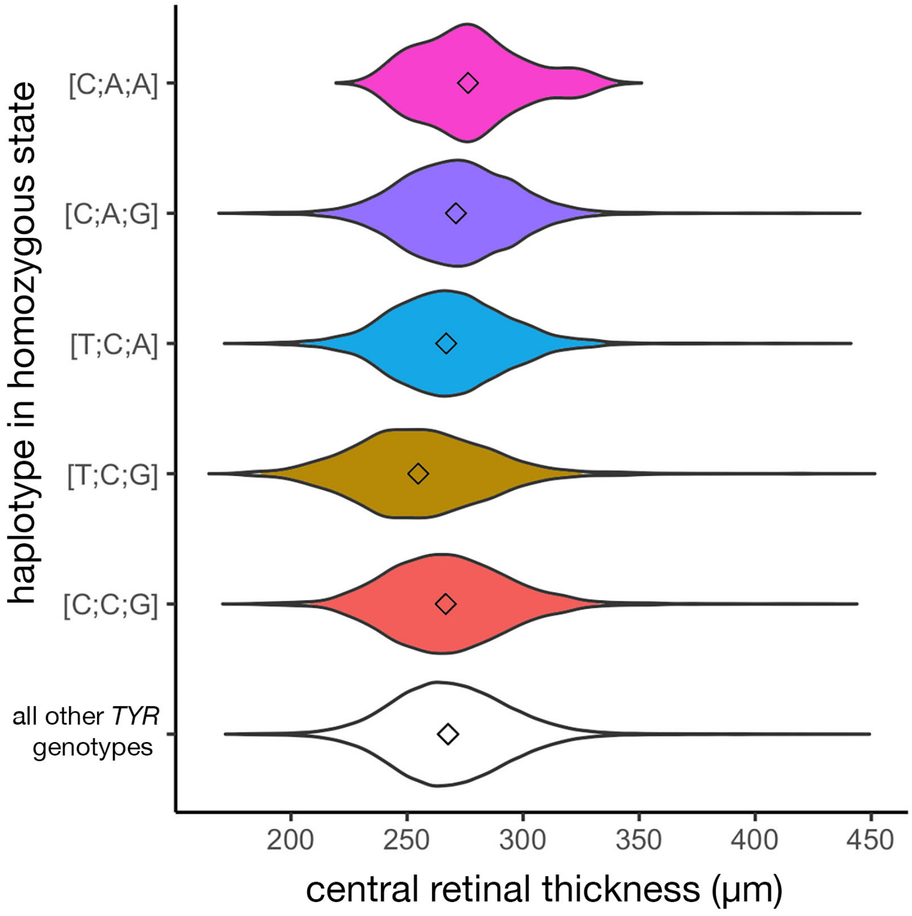


The distribution of central retinal thickness in UK Biobank participants who are homozygous for various *TYR* haplotypes is shown. The Kruskal-Wallis p-value was $< 2 \times 10^{-16}$ and further information including numeric data can be found in Supplementary Table 6.

[C;A;A] corresponds to homozygosity for *TYR* c.[-301C;575A;1205A], a genotype found to confer a high albinism risk;
[C;A;G] corresponds to homozygosity for *TYR* c.[-301C;575A;1205G];
[T;C;A] corresponds to homozygosity for *TYR* c.[-301T;575C;1205A], a genotype found to protect against albinism;
[T;C;G] corresponds to homozygosity for *TYR* c.[-301T;575C;1205G], a genotype found to protect against albinism;
[C;C;G] corresponds to homozygosity for *TYR* c.[-301C;575C;1205G].
It is noted that people with albinism tend to have increased central retinal thickness (due to underdevelopment of the fovea).

**Supplementary Figure 3**. Secondary analyses in selected subsets of the case and control cohorts confirm that common *TYR* variants form haplotypes that affect albinism risk

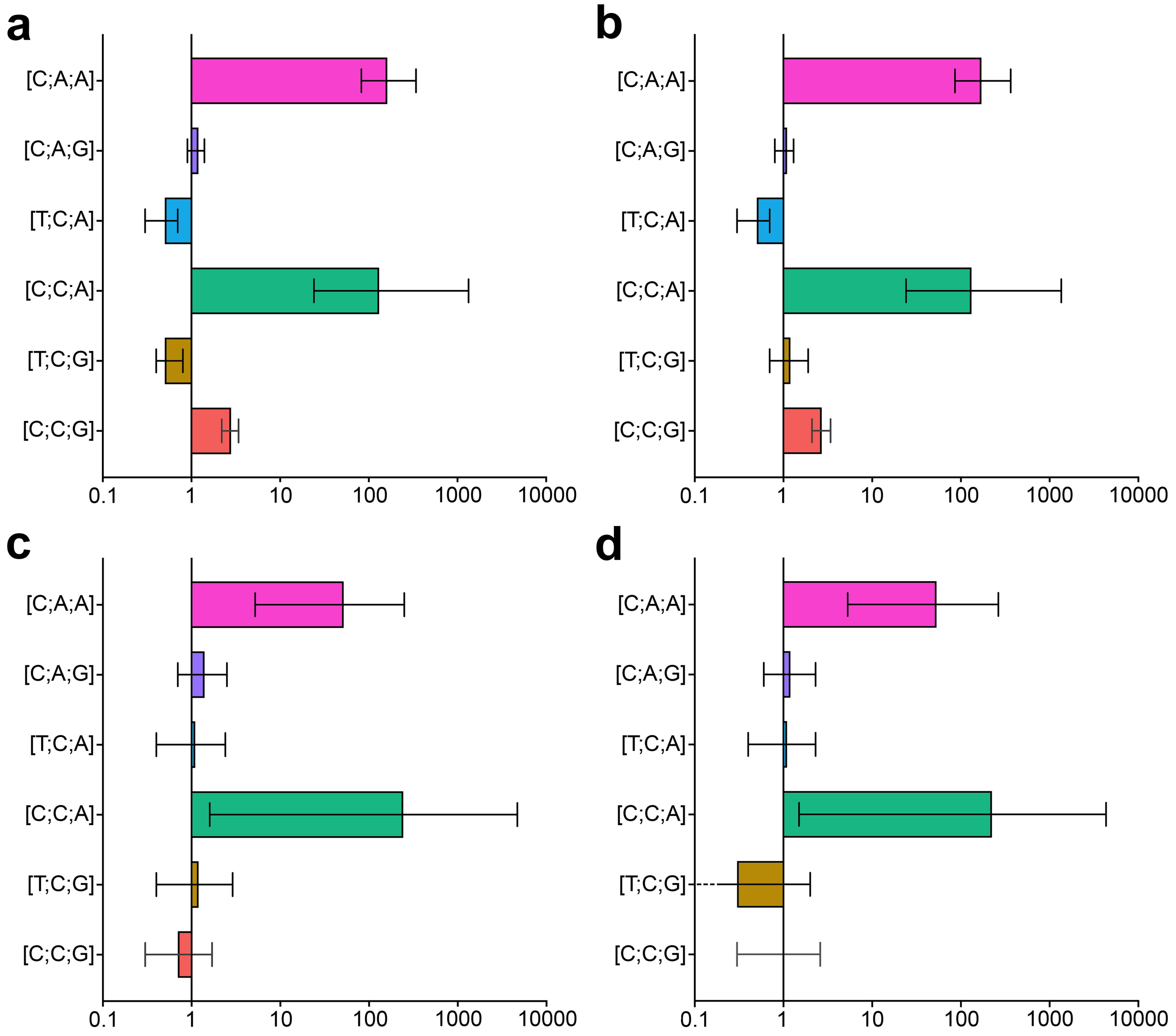


The x-axes are showing odds ratios ($log_{10}$) and correspond to the probability of receiving a diagnosis of albinism in individuals who carry a specific haplotype is homozygous state.

Panel **a**, shows the results obtained following analysis of a mixed case cohort (including 1208 probands from the University Hospital of Bordeaux cohort and 105 cases from the 100K_GP dataset) and an inclusive control cohort (29,497 unrelated individuals from the 100K_GP dataset). This graph is identical to that shown in Figure 2b and is included here to facilitate comparison with the graphs in the other panels. Further information and relevant numeric data can be found in Supplementary Table 3.

Panel **b**, shows the results obtained following a similar analysis to that in panel **a**, but this time only focusing on individuals with predominantly European ancestries. Further information and relevant numeric data can be found in Supplementary Table 10.

Panel **c** shows the results obtained following analysis of cases and controls from the 100K_GP dataset only. Further information and relevant numeric data can be found in Supplementary Table 8.

Panel **d** shows the results obtained following a similar analysis to that in panel **c**, but this time only focusing on individuals with predominantly European ancestries. Further information and relevant numeric data can be found in Supplementary Table 9.

Overall, the findings of the sub-analyses discussed in panels **b, c** and **d** are well aligned with those of the primary analysis shown in panel **a** (and Figure 2b)

[C;A;A] corresponds to homozygosity for *TYR* c.[-301C;575A;1205A];
[C;A;G] corresponds to homozygosity for *TYR* c.[-301C;575A;1205G];
[T;C;A] corresponds to homozygosity for *TYR* c.[-301T;575C;1205A];
[C;C;A] corresponds to homozygosity for *TYR* c.[-301C;575C;1205A];
[T;C;G] corresponds to homozygosity for *TYR* c.[-301T;575C;1205G];
[C;C;G] corresponds to homozygosity for *TYR* c.[-301C;575C;1205G];
100K_GP corresponds to Genomics England 100,000 Genomes Project.

#### SUPPLEMENTARY TABLES

**Supplementary Table 1**. Findings in 1,208 probands with albinism who underwent clinical-grade genetic testing at the University Hospital of Bordeaux Molecular Genetics Laboratory.

(see spreadsheet)

**Supplementary Table 2**. Cohort characteristics

| | **cases** | **controls** |
|---|---|---|
| number of probands | 1,208 + 105 [a] | 29,497 |
| percentage of female probands | 44% [b] | 48% |
| percentage of probands with European ancestry [c] | 82% | 78% |
| genetic testing approach | gene panel testing or genome sequencing [a] | genome sequencing |
| probability of a proband carrying two or more rare HGMD-listed alleles [d] in the same albinism-associated gene | 38% | 0.4% |

[a] 1,208 probands with a diagnosis of albinism were identified through the University Hospital of Bordeaux Molecular Genetics Laboratory database; most of these cases underwent gene panel testing. A further 105 affected probands were identified in the Genomics England 100,000 Genomes Project dataset; these cases underwent genome sequencing using a similar pipeline to the control cohort.

[b] further analysis revealed that the observed enrichment in male probands in the case cohort was due to the contribution of X-linked forms of albinism.

[c] ancestry was estimated using a questionnaire in most affected probands and using principal component analysis in the control cohort (see Methods).

[d] refers to single nucleotide variants (or small insertions/deletions) that have minor-allele frequency [MAF] < 1% and are labelled as disease-causing (DM) in the Human Gene Mutation Database (HGMD) v2021.2.

**Supplementary Table 3.** Risk of albinism in people carrying selected *TYR* haplotypes in homozygous state

| ***TYR* haplotype (in homozygous state)** | | | **odds ratio** [b] | **95% confidence interval** | **p-value** |
|---|---|---|---|---|---|
| c.-301C>T [rs4547091] [a] | c.575C>A [rs1042602] | c.1205G>A [rs1126809] | | | |
| C (↓) | A | A | 162 | 82-341 | <0.00001 |
| C (↓) | A | G | 1.2 | 0.9-1.4 | 0.16 |
| T (↑) | A | A | genotype not detected [c] | | |
| T (↑) | A | G | genotype not detected [c] | | |
| T (↑) | C | A | 0.5 | 0.3-0.7 | 0.00003 |
| C (↓) | C | A | 131 | 24-1336 | <0.00001 |
| T (↑) | C | G | 0.5 | 0.4-0.8 | 0.0007 |
| C (↓) | C | G | 2.8 | 2.2-3.4 | <0.00001 |
| **covariates** | | | | | |
| male gender | | | 1.3 | 1.2-1.5 | 0.00002 |
| presence of one rare risk genotype [d] | | | 7.0 | 6.0-8.1 | <0.00001 |
| presence of two or more rare risk genotypes [d] | | | 100 | 83-121 | <0.00001 |
| ancestry [e] (African) | | | 4.1 | 2.9-5.5 | <0.00001 |
| ancestry [e] (American) | | | 2.5 | 1.2-4.8 | 0.02 |
| ancestry [e] (East Asian) | | | 0.2 | 0.04-0.6 | 0.003 |
| ancestry [e] (South Asian) | | | 0.1 | 0.1-0.2 | <0.00001 |

[a] the reference allele of this promoter variant (c.-301C) reduces *TYR* expression and is shown as C (↓); the non-reference allele (c.-301T) increases gene expression and is shown as T (↑).

[b] Firth logistic regression was used to generate odds ratios and p-values.

[c] the [T;A;A] and [T;A;G] haplotypes were not detected in homozygous state in neither the case nor the control cohort; this is likely to be due to the strong linkage disequilibrium between the *TYR* c.-301T and c.575C alleles in European populations (D' 1.0, $R^2$ 0.35 in British populations included in the 1000 Genomes Project[1]).

[d] one risk genotype corresponds to a heterozygous single nucleotide variant (or small insertion/deletion) with a minor-allele frequency [MAF] < 1% and a "disease-causing" (DM) label in the Human Gene Mutation Database (HGMD) v2021.2; only variants in autosomal albinism-associated genes were considered.

[e] ancestry was estimated using a questionnaire in most affected probands and using principal component analysis in the control cohort (see Methods).

A plot of these results can be found in Figure 2b and in Supplementary Figure 3a.

**Supplementary Table 4**. Differences in the diagnostic yield of genetic testing and the prevalence of *TYR*-associated albinism between individuals with presumed European and presumed African ancestries.

| | number of affected probands in the University Hospital of Bordeaux cohort | percentage of probands with a molecular diagnosis of albinism following clinical-grade genetic testing [a] | percentage of probands with a molecular diagnosis of *TYR*-associated albinism following clinical-grade genetic testing [b] | percentage of individuals carrying the c.-301C>T [rs4547091] genotype [c] |
|---|---|---|---|---|
| individuals with albinism and presumed European ancestries | 1,032 | 70% | 34% | 30% (heterozygous; CT)<br>7% (homozygous; TT) |
| individuals with albinism and presumed African ancestries | 156 | 89% | 13% | 17% (heterozygous; CT)<br>32% (homozygous; TT) |

[a] Clinical-grade genetic testing refers to the genetic analysis that was performed at the University Hospital of Bordeaux Molecular Genetics Laboratory. This involved analyzing single-nucleotide changes, copy-number changes and the high-risk genotypes reported here (*i.e.* homozygosity for the c.[-301C;575A;1205A] and c.[-301C;575C;1205A] *TYR* haplotypes).

[b] The difference in the prevalence of *TYR*-associated albinism between people with presumed European and people with presumed African ancestries was statistically significant (chi-squared p-value < 0.00001).

[c] The non-reference allele of the *TYR* promoter variant c.-301C>T increases gene expression. The higher prevalence of TT genotypes in people with presumed African ancestries may partly account for the relatively low prevalence of *TYR*-associated albinism in these populations.

**Supplementary Table 5.** Visual acuity in UK Biobank participants carrying selected *TYR* haplotypes in homozygous state.

| ***TYR* haplotype (in homozygous state)** | | | **Number of people with visual acuity data in the UK Biobank** | **Median LogMAR visual acuity** | **Mean LogMAR visual acuity** |
|---|---|---|---|---|---|
| c.-301C>T [rs4547091] | c.575C>A [rs1042602] | c.1205G>A [rs1126809] | | | |
| C (↓) | A | A | 33 | 0.06 | 0.10 |
| C (↓) | A | G | 12,173 | -0.01 | 0.03 |
| T (↑) | A | A | 0 | not applicable | |
| T (↑) | A | G | 0 | not applicable | |
| T (↑) | C | A | 7,413 | -0.01 | 0.02 |
| C (↓) | C | A | 1 | 0.49 | 0.49 |
| T (↑) | C | G | 4,824 | 0.0 | 0.04 |
| C (↓) | C | G | 6,511 | -0.01 | 0.03 |
| **all other *TYR* genotypes (including heterozygous)** | | | 79,558 | -0.01 | 0.03 |

These findings are plotted in Figure 2. Given that:

- no UK Biobank participants were homozygous for the [T;A;A] and [T;A;G] haplotypes,
- only one UK Biobank participant had visual acuity assessment and was homozygous for the [C;C;A] haplotype,

these three genotypes were excluded from further statistical analyses. The average ranks of the remaining six categories were compared using the Kruskal-Wallis method; a p-value of $8 \times 10^{-11}$ was obtained. Pair-wise comparisons between homozygosity for [C;A;A] and the other categories revealed the following Benjamini-Hochberg adjusted p-values:

[C;A;A] vs [C;A;G], p-value 0.013;
[C;A;A] vs [T;C;A], p-value 0.010;
[C;A;A] vs [T;C;G], p-value 0.039;
[C;A;A] vs [C;C;G], p-value 0.013;
[C;A;A] vs all other *TYR* genotypes, p-value 0.011.

A plot of these results can be found in Figure 2c.

**Supplementary Table 6.** Central retinal thickness in UK Biobank participants carrying selected *TYR* haplotypes in homozygous state

| ***TYR* haplotype (in homozygous state)** | | | **Number of people with optical coherence tomography (OCT) data in the UK Biobank** | **Median central retinal thickness** | **Mean central retinal thickness** |
|---|---|---|---|---|---|
| c.-301C>T [rs4547091] | c.575C>A [rs1042602] | c.1205G>A [rs1126809] | | | |
| C (↓) | A | A | 20 | 276 | 278 |
| C (↓) | A | G | 7,338 | 271 | 271 |
| T (↑) | A | A | 0 | not applicable | |
| T (↑) | A | G | 0 | not applicable | |
| T (↑) | C | A | 4, 553 | 267 | 268 |
| C (↓) | C | A | 0 | not applicable | |
| T (↑) | C | G | 2,674 | 255 | 256 |
| C (↓) | C | G | 3,782 | 267 | 268 |
| **all other *TYR* genotypes (including heterozygous)** | | | 45,226 | 268 | 269 |

These findings are plotted in Supplementary Figure 2. Given that no UK Biobank participants had OCT imaging of sufficient quality and were homozygous for the [C;C;A], [T;A;A] or [T;A;G] haplotypes, these three genotypes were excluded from further statistical analyses. The average ranks of the remaining six categories were compared using the Kruskal-Wallis method; a p-value of $< 2 \times 10^{-16}$ was obtained. Pair-wise comparisons between homozygosity for [C;A;A] and the other categories revealed the following Benjamini-Hochberg adjusted p-values:
[C;A;A] vs [C;A;G], p-value 0.13;
[C;A;A] vs [T;C;A], p-value 0.10;
[C;A;A] vs [T;C;G], p-value 0.0007;
[C;A;A] vs [C;C;G], p-value 0.10;
[C;A;A] vs all other *TYR* genotypes, p-value 0.13.

A plot of these results can be found in Supplementary Figure 2.

**Supplementary Table 7**. Single-nucleotide variants used to calculate the genomic inflation factor lambda (λGC).

| variant coordinates (GRCh38) | variant dbSNP rsID | variant consequence (MANE select transcript) | name of associated gene (HGNC) | $\chi^2$ (Firth regression) | gnomAD v3.1.2 frequency in NFE | CADD Phred score | SpliceAI score |
|---|---|---|---|---|---|---|---|
| 12-88507004-A-T | rs183924903 | ENST00000644744.1: c.714+24T>A | *KITLG* | 0.099 | 0.001705 | 0.002 | 0 |
| 3-52404411-C-T | rs150454901 | ENST00000460680.6: c.1250+42G>A | *BAP1* | 0.976 | 0.003101 | 0.017 | 0 |
| 3-119490496-A-G | rs142663532 | ENST00000295588.9: c.798-55A>G | *POGLUT1* | 0.314 | 0.00369 | 0.041 | 0 |
| 16-84016604-G-A | rs77876966 | ENST00000299709.8: p.Leu359Leu | *SLC38A8* | 0.864 | 0.05285 | 0.075 | 0.02 |
| 5-33944723-G-A | rs150473213 | ENST00000296589.9: p.Val506Val | *SLC45A2* | 9.828 | 0.0002205 | 0.077 | 0 |
| 15-58646011-A-G | rs201948093 | ENST00000260408.8: c.735+44T>C | *ADAM10* | 0.18 | 0.003235 | 0.191 | 0 |
| 6-397261-G-A | rs34318727 | ENST00000380956.9: c.637+9G>A | *IRF4* | 16.21 | 0.07294 | 0.193 | 0 |
| 3-133953627-T-C | rs199782188 | ENST00000310926.11: c.724+36A>G | *SLCO2A1* | 0.042 | 0.01306 | 0.222 | 0 |
| 1-154589737-T-G | rs115805812 | ENST00000368474.9: c.2668+20A>C | *ADAR* | 0.039 | 0.004735 | 0.247 | 0 |
| 10-98429811-C-A | rs11592273 | ENST00000361490.9: p.Gly283Trp | *HPS1* | 1.937 | 0.0758 | 0.337 | 0 |
| 15-58646039-G-A | rs138743329 | ENST00000260408.8: c.735+16C>T | *ADAM10* | 0.409 | 0.00178 | 0.385 | 0 |
| 3-69949177-C-G | rs181810413 | ENST00000352241.9: c.880+9C>G | *MITF* | 0.414 | 0.003381 | 0.553 | 0 |
| 15-55228733-T-C | rs368760836 | ENST00000336787.6: c.240-21A>G | *RAB27A* | 2.213 | 0.0001176 | 0.611 | 0 |
| 20-32230808-G-A | rs200896099 | ENST00000375749.8: c.736-11G>A | *POFUT1* | 0.143 | 0.001911 | 0.685 | 0.09 |
| 19-2114320-G-A | rs117387599 | ENST00000643116.3: c.2424-18C>T | *AP3D1* | 2.605 | 0.01143 | 0.698 | 0 |
| 15-27871136-T-C | rs41304383 | ENST00000354638.8 : c.2244+18A>G | *OCA2* | 5.378 | 0.04011 | 0.831 | 0 |
| 4-54709427-C-T | rs72549293 | ENST00000288135.6: p.Tyr373Tyr | *KIT* | 0.983 | 0.0001029 | 0.894 | 0.03 |
| 12-52516840-G-A | rs144359915 | ENST00000252242.9: p.Asn412Asn | *KRT5* | 0.021 | 0.002307 | 0.94 | 0 |
| 9-21974641-C-G | rs45456595 | ENST00000304494.9: c.150+37G>C | *CDKN2A* | 0.177 | 0.003248 | 1.22 | 0.29 |
| 1-154597286-C-T | rs200651742 | ENST00000368474.9: c.1935-19G>A | *ADAR* | 0.069 | 0.00172 | 1.26 | 0 |
| 6-15524467-T-C | rs16876573 | ENST00000344537.10: c.811+59A>G | *DTNBP1* | 0.472 | 0.04261 | 1.9 | 0 |
| 4-174492145-C-T | rs17060532 | ENST00000296522.11: c.663-51G>A | *HPGD* | 0.016 | 0.04114 | 2.05 | 0 |
| (continued) | | | | | | | |

| variant coordinates (GRCh38) | variant dbSNP rsID | variant consequence (MANE select transcript) | name of associated gene (HGNC) | $\chi^2$ (Firth regression) | gnomAD v3.1.2 frequency in NFE | CADD Phred score | SpliceAI score |
|---|---|---|---|---|---|---|---|
| 6-131851119-A-G | rs200746177 | ENST00000647893.1: c.431-23A>G | *ENPP1* | 0.375 | 0.001323 | 2.26 | 0 |
| 6-15533192-C-A | rs75380691 | ENST00000344537.10: c.667+48G>T | *DTNBP1* | 1.93 | 0.005808 | 2.69 | 0.04 |
| 12-88506969-A-G | rs41283110 | ENST00000644744.1: c.714+59T>C | *KITLG* | 4.972 | 0.007056 | 2.76 | 0 |
| 2-219212539-T-C | rs114354921 | ENST00000265316.9: c.1864-48A>G | *ABCB6* | 0.039 | 0.005657 | 3.11 | 0 |
| 15-52372211-G-A | rs11637651 | ENST00000399233.6: p.Ile910Ile | *MYO5A* | 0.892 | 0.06795 | 3.24 | 0 |
| 12-52519946-G-A | rs11549951 | ENST00000252242.9: p.Leu117Leu | *KRT5* | 0.359 | 0.07463 | 3.29 | 0 |
| 12-52517611-G-A | rs149467228 | ENST00000252242.9: p.Ala357Ala | *KRT5* | 1.656 | 0.007071 | 3.56 | 0.01 |
| 19-2113468-G-A | rs146083008 | ENST00000643116.3: c.2602-55C>T | *AP3D1* | 0.029 | 0.02771 | 3.83 | 0.06 |
| 3-149155185-G-A | rs34197730 | ENST00000296051.7: p.Thr493Thr | *HPS3* | 0.601 | 0.03576 | 4.28 | 0.13 |
| 12-57751138-C-G | rs3211614 | ENST00000257904.11: c.355-48G>C | *CDK4* | 1.646 | 0.002073 | 4.51 | 0.06 |
| 12-88516544-G-T | rs182948635 | ENST00000644744.1: c.364-54C>A | *KITLG* | 0.02 | 0.006951 | 4.53 | 0 |
| 2-237540465G-T | rs61737681 | ENST00000264605.8: p.Ala408Ser | *MLPH* | 1E+17 | 0.03652 | 0.116 | 0 |
| 10-102065990-G-A | rs3737243 | ENST00000299238.7: p.Gly172Gly | *HPS6* | 6.705 | 0.1081 | 4.59 | 0 |

gnomAD v3.1.2 frequency in NFE corresponds to the minor allele frequency in the non-Finnish European subset of the Genome Aggregation Database (gnomAD) v3.1.2. We selected variants with an allele frequency broadly matching this of the genotypes studied in the case-control analysis.

CADD corresponds to Combined Annotation-Dependent Depletion, an integrative annotation tool for genetic variants. CADD provides a ranking rather than a prediction and the PHRED-scaled score ranges from 1 to 99 (with higher values indicating more deleterious cases). For the λGC analysis we wanted to select variants that are unlikely to affect protein function and we only chose changes with a CADD PHRED-scaled score less than 5.

It is noted that $\lambda_{median}$ was found to be 1.04. Given that this is close to the expected value of 1.00, we can conclude that there is limited evidence of confounding by ancestry.

**Supplementary Table 8.** Risk of albinism in people carrying selected *TYR* haplotypes in homozygous state: sub-analysis of cases and controls from the Genomics England 100,000 Genomes Project cohort

| ***TYR* haplotype (in homozygous state)** | | | **odds ratio** [b] | **95% confidence interval** | **p-value** |
|---|---|---|---|---|---|
| c.-301C>T [rs4547091] [a] | c.575C>A [rs1042602] | c.1205G>A [rs1126809] | | | |
| C (↓) | A | A | 52 | 5.2-250 | 0.0036 |
| C (↓) | A | G | 1.4 | 0.7-2.5 | 0.3 |
| T (↑) | C | A | 1.1 | 0.4-2.4 | 0.8 |
| C (↓) | C | A | 243 | 1.7-4693 | 0.037 |
| T (↑) | C | G | 1.2 | 0.4-2.9 | 0.8 |
| C (↓) | C | G | 0.7 | 0.3-1.7 | 0.5 |
| **covariates** | | | | | |
| male gender | | | 0.9 | 0.6-1.3 | 0.6 |
| presence of one rare risk genotype [c] | | | 6.3 | 3.9-10 | <0.00001 |
| presence of two or more rare risk genotypes [c] | | | 60 | 35-101 | <0.00001 |
| ancestry [d] (African) | | | 1.5 | 0.5-4.4 | 0.5 |
| ancestry [d] (American) | | | 5.9 | 0.7-23 | 0.1 |
| ancestry [d] (East Asian) | | | 1.1 | 0.01-8.4 | 0.9 |
| ancestry [d] (South Asian) | | | 1.8 | 0.9-3.2 | 0.1 |

[a] the reference allele of this promoter variant (c.-301C) reduces *TYR* expression and is shown as C (↓); the non-reference allele (c.-301T) increases gene expression and is shown as T (↑).

[b] Firth logistic regression was used to generate odds ratios and p-values.

[c] one risk genotype corresponds to a heterozygous single nucleotide variant (or small insertion/deletion) with a minor-allele frequency [MAF] < 1% and a "disease-causing" (DM) label in the Human Gene Mutation Database (HGMD) v2021.2; only variants in autosomal albinism-associated genes were considered.

[d] ancestry was inferred using principal component analysis and five broad super-populations were projected (European, African, Admixed American, East Asian, South Asian) (see Methods).

A plot of these results can be found in Supplementary Figure 3b.

**Supplementary Table 9**. Risk of albinism in people carrying selected *TYR* haplotypes in homozygous state: sub-analysis of cases and controls from the Genomics England 100,000 Genomes Project cohort (European subset)[a].

| ***TYR* haplotype (in homozygous state)** | | | **odds ratio[c]** | **95% confidence interval** | **p-value** |
|---|---|---|---|---|---|
| c.-301C>T [rs4547091][b] | c.575C>A [rs1042602] | c.1205G>A [rs1126809] | | | |
| C (↓) | A | A | 53 | 5.3-264 | 0.0035 |
| C (↓) | A | G | 1.2 | 0.6-2.3 | 0.56 |
| T (↑) | A | A | genotype not detected[c] | | |
| T (↑) | A | G | genotype not detected[c] | | |
| T (↑) | C | A | 1.1 | 0.4-2.3 | 0.9 |
| C (↓) | C | A | 224 | 1.5-4332 | 0.04 |
| T (↑) | C | G | 0.3 | 0.001-2 | 0.2 |
| C (↓) | C | G | 0.9 | 0.3-2.6 | 0.99 |
| **covariates** | | | | | |
| male gender | | | 0.9 | 0.6-1.4 | 0.6 |
| presence of one rare risk genotype[d] | | | 6 | 3.5-10 | <0.00001 |
| presence of two or more rare risk genotypes[d] | | | 50 | 27-92 | <0.00001 |

[a] ancestry was inferred in cases and controls from the Genomics England 100,000 Genomes Project cohort using principal component analysis; only probands matching to the European subset of the 1000 genomes project (phase 3) dataset were included in this analysis (see Methods).

[b] the reference allele of this promoter variant (c.-301C) reduces *TYR* expression and is shown as C (↓); the non-reference allele (c.-301T) increases gene expression and is shown as T (↑).

[c] Firth logistic regression was used to generate odds ratios and p-values.

[d] one risk genotype corresponds to a heterozygous single nucleotide variant (or small insertion/deletion) with a minor-allele frequency [MAF] < 1% and a “disease-causing” (DM) label in the Human Gene Mutation Database (HGMD) v2021.2; only variants in autosomal albinism-associated genes were considered.

A plot of these results can be found in Supplementary Figure 3c.

**Supplementary Table 10.** Risk of albinism in people carrying selected *TYR* haplotypes in homozygous state: sub-analysis of cases and controls from the University Hospital of Bordeaux and the Genomics England 100,000 Genomes Project cohorts (European subset) [a]

| ***TYR* haplotype (in homozygous state)** | | | **odds ratio** [b] | **95% confidence interval** | **p-value** |
|---|---|---|---|---|---|
| c.-301C>T [rs4547091] [a] | c.575C>A [rs1042602] | c.1205G>A [rs1126809] | | | |
| C (↓) | A | A | 170 | 86-365 | <0.00001 |
| C (↓) | A | G | 1.1 | 0.8-1.3 | 0.6 |
| T (↑) | A | A | genotype not detected [c] | | |
| T (↑) | A | G | genotype not detected [c] | | |
| T (↑) | C | A | 0.5 | 0.3-0.7 | 0.00003 |
| C (↓) | C | A | 132 | 24-1353 | <0.00001 |
| T (↑) | C | G | 1.2 | 0.7-1.9 | 0.4 |
| C (↓) | C | G | 2.7 | 2.1-3.4 | <0.00001 |
| **covariates** | | | | | |
| male gender | | | 1.4 | 1.2-1.6 | <0.00001 |
| presence of one rare risk genotype [d] | | | 7.5 | 6.4-8.7 | <0.00001 |
| presence of two or more rare risk genotypes [d] | | | 101 | 82-123 | <0.00001 |

[a] ancestry was estimated using a questionnaire in most cases (1,208/1,313) and using principal component analysis in the remaining cases and in controls (see Methods).

[b] the reference allele of this promoter variant (c.-301C) reduces *TYR* expression and is shown as C (↓); the non-reference allele (c.-301T) increases gene expression and is shown as T (↑).

[c] Firth logistic regression was used to generate odds ratios and p-values.

[d] one risk genotype corresponds to a heterozygous single nucleotide variant (or small insertion/deletion) with a minor-allele frequency [MAF] < 1% and a "disease-causing" (DM) label in the Human Gene Mutation Database (HGMD) v2021.2; only variants in autosomal albinism-associated genes were considered.

A plot of these results can be found in Supplementary Figure 3d.

#### REFERENCES


1. Auton, A. *et al.* A global reference for human genetic variation. *Nature* **526**, 68–74 (2015).

2. Alexander, T. A. & Machiela, M. J. LDpop: an interactive online tool to calculate and visualize geographic LD patterns. *BMC Bioinformatics* **21**, 1–4 (2020).

3. Mallick, S. *et al.* The Simons Genome Diversity Project: 300 genomes from 142 diverse populations. *Nat. 2016 5387624* **538**, 201–206 (2016).

4. Albers, P. K. & McVean, G. Dating genomic variants and shared ancestry in population-scale sequencing data. *PLOS Biol.* **18**, e3000586 (2020).

5. Hudjashov, G., Villems, R. & Kivisild, T. Global Patterns of Diversity and Selection in Human Tyrosinase Gene. *PLoS One* **8**, e74307 (2013).

6. Brownrigg, R., Minka, T. P., Deckmyn, A., Becker, R. A. & Wilks, A. R. maps: Draw geographical maps R package version 3.4.0. (2021).

7. Lemon, J. *et al.* plotrix: Plotting functions R package version 3.8-2. (2021).
